# Prevalence and predictors of Motoric Cognitive Risk syndrome in a community-dwelling older Scottish population: a longitudinal observational study

**DOI:** 10.1101/2022.07.21.22277916

**Authors:** Donncha S. Mullin, Lucy E Stirland, Miles Welstead, Tom C. Russ, Michelle Luciano, Graciela Muniz-Terrera

## Abstract

**Objectives:** This study aimed to determine the prevalence of Motoric Cognitive Risk (MCR), describe associated risk factors for this syndrome, and assess its overlap with Mild Cognitive Impairment, Prefrailty, and Frailty, in a cohort of older Scottish adults.

**Methods:** A longitudinal prospective study using data from the Lothian Birth Cohort 1936 (LBC1936) and follow-up data from six, nine, and 12 years later. A total of 690 participants (mean [SD] age 76.3 [0.8] years) free from dementia were classified into non-MCR or MCR groups and baseline characteristics (age 69.5 [0.8] years) between the groups were compared.

**Results:** MCR prevalence rate ranged from 5.3-5.7% across the three waves. The presence of MCR was significantly associated with older age (six and nine years later), lower occupational socioeconomic status (six years later), and a range of tests of executive function (six, nine and 12 years later). Approximately 46% of the MCR group also had Mild Cognitive Impairment and almost all the MCR group had either Prefrailty or Frailty.

**Conclusions:** The prevalence of MCR in this Scottish cohort is lower than the pooled global average but higher than the prevalence in neighbouring countries. Future LBC1936 research should assess the risk factors associated with MCR to validate previous findings and analyse novel predictive factors, particularly socioeconomic status. This study can serve as a foundation for future studies to improve dementia risk assessments and potentially develop new interventions to reduce incident dementia.

**Key points:**

1. Motoric Cognitive Risk (MCR) is a gait-based predementia syndrome that is quick, inexpensive, and practical to assess and diagnose, and it can identify individuals at high risk of developing dementia
2. The prevalence of MCR in this older Scottish cohort ranged from 5.3-5.7% over three follow-up waves
3. Factors associated with MCR in this cohort include age, socioeconomic status and tests of executive function
4. There is partial overlap between individuals with MCR and Mild Cognitive Impairment (MCI), but almost all individuals with MCR also had either Prefrailty or Frailty.

## Introduction

Dementia is a leading cause of morbidity and mortality globally.^1^ Effective treatments for dementia remain elusive. Any method that supports the early identification of high-risk individuals would allow for addressing modifiable risk factors and organising future care needs. This method would also assist research trials with cohort recruitment and ultimately contribute to a reduction in the prevalence of dementia. The Motoric Cognitive Risk (MCR) syndrome is a high-risk state combining objective (measured) slow walking speed and subjective (self-reported) cognitive complaint in the absence of significant functional impairment and dementia.^2^ Slow gait speed and subjective cognitive complaints are some of the earliest reported findings in the pre-clinical stage of dementia, occurring approximately 10 years before dementia diagnosis.^3^

MCR is quick, inexpensive, and practical to assess and diagnose. It does not require any expensive technology, specialised assessment, invasive investigations, or brain imaging scans. Thus, MCR could be useful in low- and middle-income countries, where currently two-thirds of the global population with dementia reside,^1^ while also having the potential to be an adjunct to memory services referrals in more economically developed countries

MCR is a recently-defined construct, first appearing in the literature in 2013^2^. As such, and despite a growing body of literature on MCR, it is still important to determine its prevalence and associated factors in diverse global populations. To date, prevalence rates range from 1.7%^4^ (Australia) to 27% (India).^5^ Generally, higher-income countries have lower prevalence rates of MCR, although how the MCR criteria are operationalised across studies also affects rates.^6^ An increasing body of work supports the prognostic utility of MCR. A 2022 systematic review and meta-analysis found that, compared to those without MCR, those with MCR are at over twice the risk of developing dementia after 4.3 years of follow-up and 76% more likely to develop cognitive impairment after 5.6 years of follow up.^7^ MCR is also prognostic of future falls and earlier mortality.^7^

This is the first study to derive MCR in a Scottish cohort, determine its prevalence and describe associated risk factors. In doing so, it is the first study to report on slow gait cut-scores in this population. Another novelty is that this study explores the overlap between MCR and the other high-risk states of Mild Cognitive Impairment (MCI), Prefrailty, and Frailty.

The aims of this study are to determine the prevalence of MCR syndrome, describe associated risk factors, and assess its overlap with Mild Cognitive Impairment, Prefrailty, and Frailty, in a cohort of older Scottish adults.

## Methods

### Study Design

This longitudinal prospective study used data from the Lothian Birth Cohort 1936 (LBC1936), which has been described in detail elsewhere.^8,9^ In summary, participants living in the Lothian region of Scotland (which includes Edinburgh), most of whom had completed an intelligence test aged 11 years, were recruited in 2004, at mean age 69.5 years (N=1091). They have been followed up every three years since, at mean ages 72.5 years (n=866), 76.3 years (n=697), 79.3 years (n=550) and 82 years (n=431). All participants are white, and the sex split is approximately equal. At each wave, participants undergo interviews, questionnaires, blood tests and physical measures, including a timed gait test over six metres. LBC1936 is conducted according to all applicable ethical guidelines and written consent is obtained from participants at each of the waves.^9^

### Participants and study size

For our analysis, we excluded participants with dementia and those missing data required to derive the MCR phenotype. We describe sample selection with reasons for dropout and exclusion given, where known, in Figure 1.

**Figure 1.**
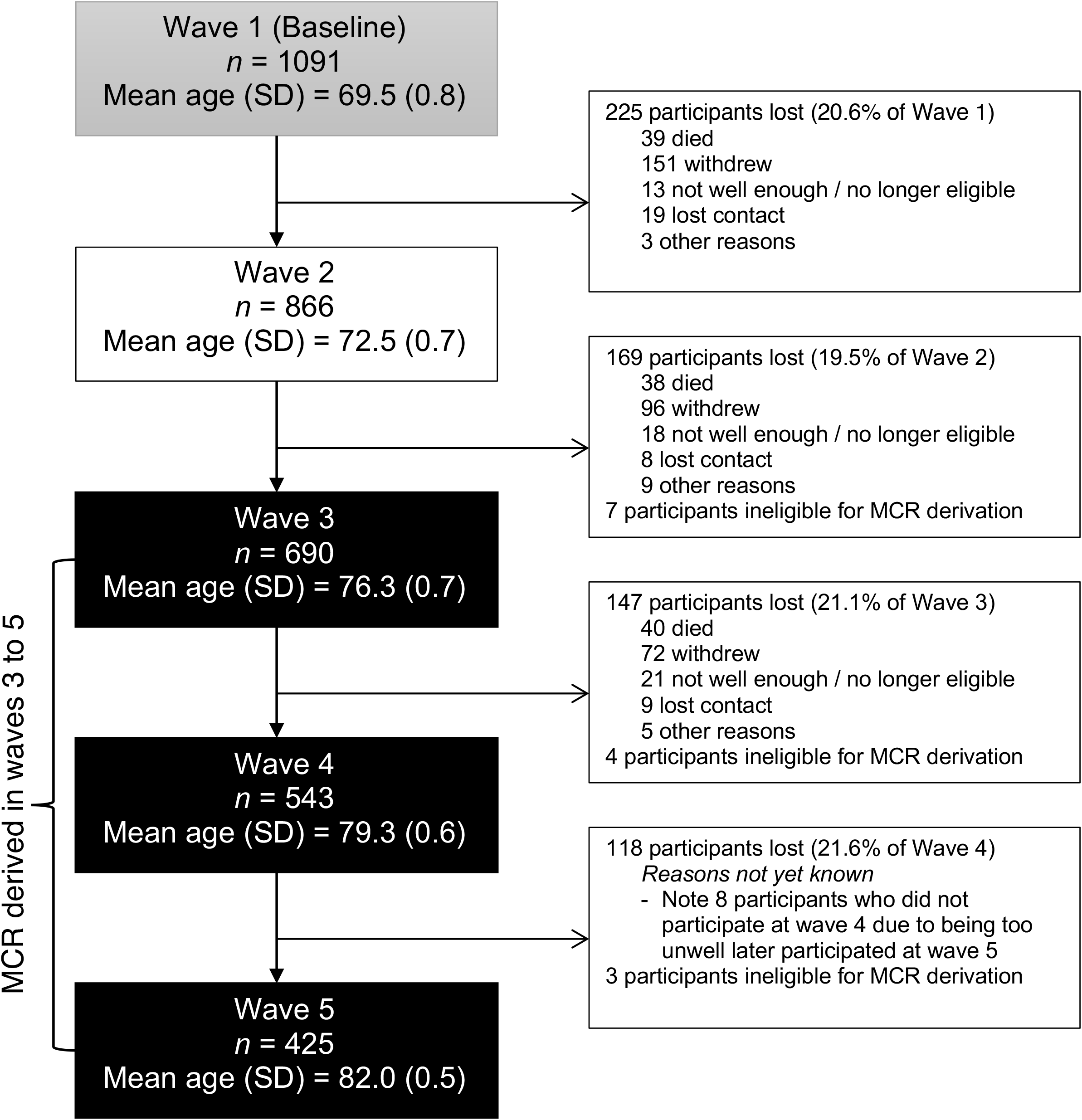
Flow chart of sample selection

### Identification of MCR

MCR is defined as the presence of subjective cognitive complaints and objective slow gait in older individuals without dementia or significant functional disability.^2^ To be classified in the MCR category, participants had to meet all four criteria reported below:

1. Slow gait as defined by walking speed one standard deviation (SD) below age- and sex-matched means. Time taken (in seconds) to walk six metres along a corridor was recorded with a stopwatch.^2^
2. Self-reported cognitive complaint: answering “yes” to the question “do you currently have any problems with your memory?”
3. No diagnosis of dementia: does not self-report a diagnosis of dementia and scores at least 24/30 on the Mini-Mental State Examination (MMSE).^10^
4. Preservation of independence in functional abilities: less than or equal to 1.5□SD above the mean on the Townsend Disability Scale overall score (a higher score indicates greater disability).^11^

We derived MCR from wave three (age 76) onwards, as the variables measuring these criteria were first identified at wave three.

### Covariates

We examined the association between the following baseline (age 69.5 [0.8] years) covariates and MCR status at waves 3, 4, and 5: age, sex, years of education, age 11 intelligence quotient (IQ; derived from the Moray House Test), marital status, body mass index (BMI), self-reported smoking status (current/ex/never), self-reported alcohol intake (units per week), depression and anxiety symptoms (Hospital Anxiety and Depression Scale), self-reported history of cardiovascular disease, and stroke. Other physical measures included forced expiratory volume in one second (FEV_1_), which is a measure of lung capacity, and grip strength (combined average of left and right). We also compared levels of blood C-reactive protein (CRP), a non-specific measure of inflammation, between non-MCR and MCR groups. All of these covariates have previously been associated with MCR^6,12,13^ except for age 11 IQ, which has never been tested. Higher childhood intelligence has previously been associated with faster gait and less subjective cognitive complaints in mid-to later-life.^14^ See **Error! Reference source not found**. for more details on how these variables were measured or derived.

### Subgroup analysis

Common to most prospective longitudinal studies of ageing, LBC1936 is susceptible to sampling bias through attrition.^8^ Compared to individuals who remained in the study, those who dropped out at each wave had lower age-11 IQ scores, lower Mini-Mental State Examination (MMSE) scores, lower socioeconomic class, and poorer physical fitness.^8^ To account for this, we performed a subgroup analysis of the MCR prevalence rates and covariates for those who withdrew compared to those who remained in the study.

### MCR overlap with MCI and Frailty levels

MCI (present/absent)^15^ and physical Frailty level according to Fried phenotype (Frail/Prefrail/not frail)^16^ have been derived in the LBC1936, as detailed in **Error! Reference source not found.**. We explored the overlap of these phenotypes with MCR within each wave of the LBC1936 dataset.

### Statistical Methods

We used descriptive analyses including the number and percentages of people with MCR to characterize the study sample. We summarised the participants’ characteristics using means and SD or frequencies and percentages, as appropriate.

We classified participants into two groups: non-MCR and MCR. These groups were compared using *χ2* tests with a continuity correction for categorical variables. For continuous explanatory variables, we performed an F-test (ANOVA) by default. We performed a Kruskal-Wallis test when variables were considered non-parametric, except in cases where Fisher’s exact test was more appropriate (i.e., when expected counts were less than five).^17^ P-values less than 0.05 were considered statistically significant. All statistical analyses were conducted in R version 4.0.2.^18^

### Missing data

We compared the distribution of all variables with missing data amongst MCR and non-MCR groups. The LBC1936 researchers have maintained a relatively low loss to follow-up rate at each wave by re-contacting those unable to attend a wave due to a temporary illness and seeing them at a later, more appropriate time where possible.^9^

## Results

Figure *1* illustrates the flow of our sample participants. We excluded three participants who had been diagnosed with dementia by the LBC1936 study doctor before wave 3. The variables necessary for deriving MCR were measured in LBC1936 from wave 3 onwards. Participants missing data in any of the necessary MCR criteria were excluded from analyses at wave 3 (*n*=4), wave 4 (*n*=4), and wave 5 (*n*=3). Accordingly, MCR status was coded for 690 participants at wave 3 (48.0% female, mean age 76.3 years), 543 participants at wave 4 (49.7% female, mean age 79.3 years), and 425 participants at wave 5 (51.1% female, mean age 82 years). Loss to follow-up in LBC1936 was approximately 20% after each wave. The main reasons for attrition were death, chronic incapacity, and permanent withdrawal.^9^ The participation rate of eligible persons was over 99% at each wave.

### MCR prevalence

MCR prevalence was very similar across waves at 5.7% (95% CI 4.0-7.6; n=39/690) at wave 3, 5.3% (95% CI 3.6-7.5; *n*=29/543) at wave 4, and 5.4% (95% CI 3.6-8.2; *n*=23/425) at wave 5. The mean prevalence of MCR was 5.5% (95% CI 4.5-6.7) across all three waves. We performed a sensitivity analysis of the MCR prevalence for those who withdrew by wave 5 (‘withdrawers’; *n*=398) compared to those who remained in the study throughout (*n*=425). MCR prevalence was higher overall amongst withdrawers at 8.6% (95% CI 5.1-11.6; *n*=23/269) at wave 3 and 11% (95% CI 5.5-16.1; *n*=14/127) at wave 4.

The gait speed cut-offs by age and sex used to define MCR are presented in Table 1.

**Table 1:** Gait speed cut-offs by age and sex in this cohort for defining Motoric Cognitive Risk.

| Age range (mean), years | Gait speed (m/s) |  |
| --- | --- | --- |
|  | Male | Female |
| 67.6 – 71.3 (69.53) | 1.36 | 1.22 |
| 71.3 – 74.6 (72.49) | 1.18 | 1.08 |
| 74.6 – 77.7 (76.24) | 1.07 | 1.00 |
| 77.7 – 80.9 (79.3) | 0.97 | 0.90 |
| 80.9 – 83.1 (82.0) | 0.96 | 0.83 |

*Note: Gait speed (m/s) values at or below the cut-off scores (one standard deviation below age- and sex-matched means) were used to define slow gait*

### Baseline covariate differences

Baseline covariate differences of the participants according to MCR status at wave 3 (six years follow-up), wave 4 (nine years follow-up) and wave 5 (12 years follow-up) are presented in Table 2. Older age was significantly associated with having MCR at waves 3 and 4, but not wave 5, despite the narrow age range of LBC1936 participants (SD 0.8 years). Sex, years of education, and age 11 IQ were not significantly associated at any wave. Lower socioeconomic status (defined by manual occupation) was significantly associated with MCR at wave 3. At all waves, poorer scores in one or more of the following tests of executive function were significantly associated with MCR compared to non-MCR: verbal fluency (wave 3), digit-symbol test (waves 3, 4 and 5), four-choice reaction time (wave 3), and block design (wave 5). The physical measures of FEV_1_ and average grip strength were significantly associated with MCR outcome at wave 4. BMI, anxiety, and depression symptoms were significantly associated with MCR at wave 5. No further covariate associations were found. There was no significant difference in missing data for any of the variables between MCR and non-MCR groups.

**Table 2:**
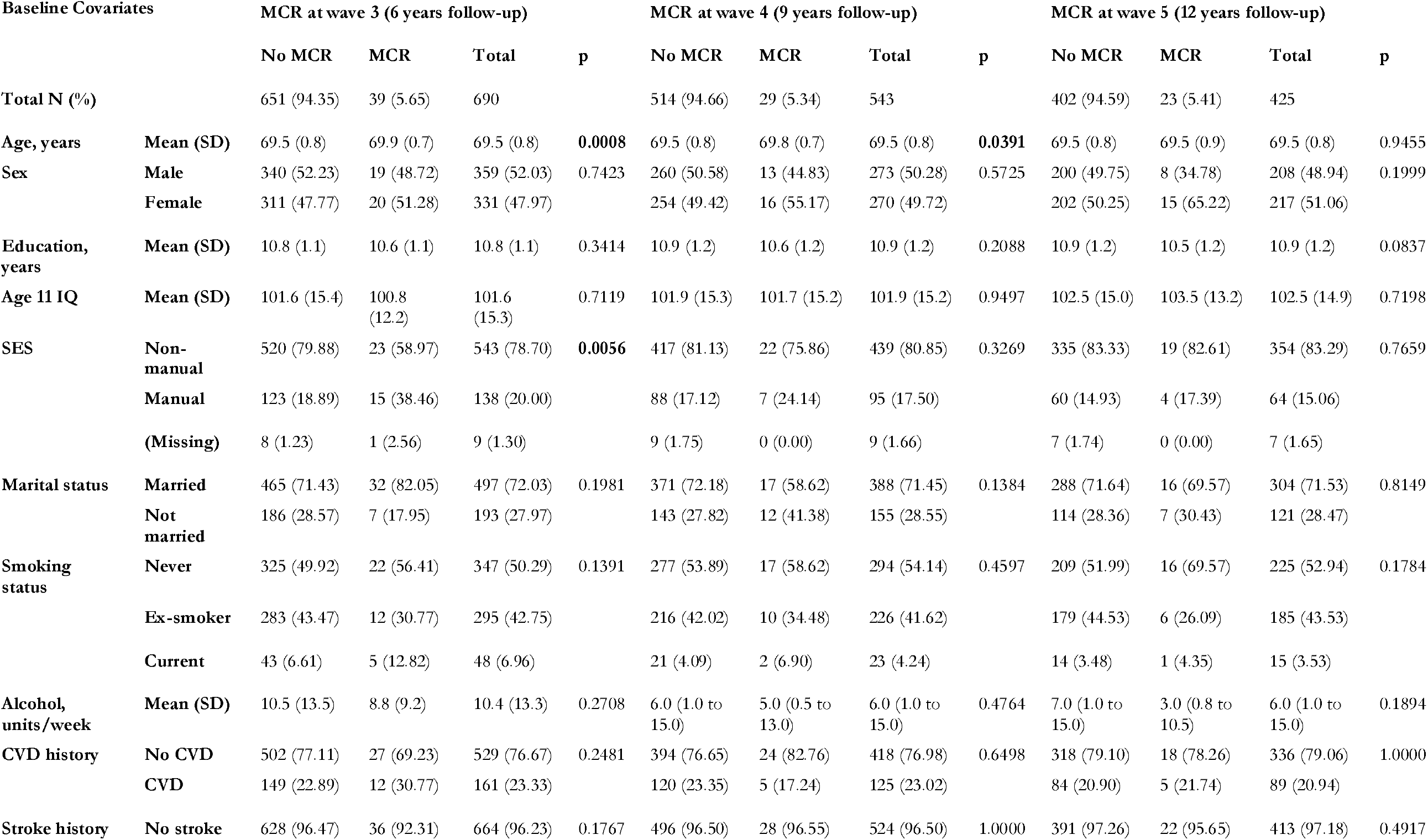
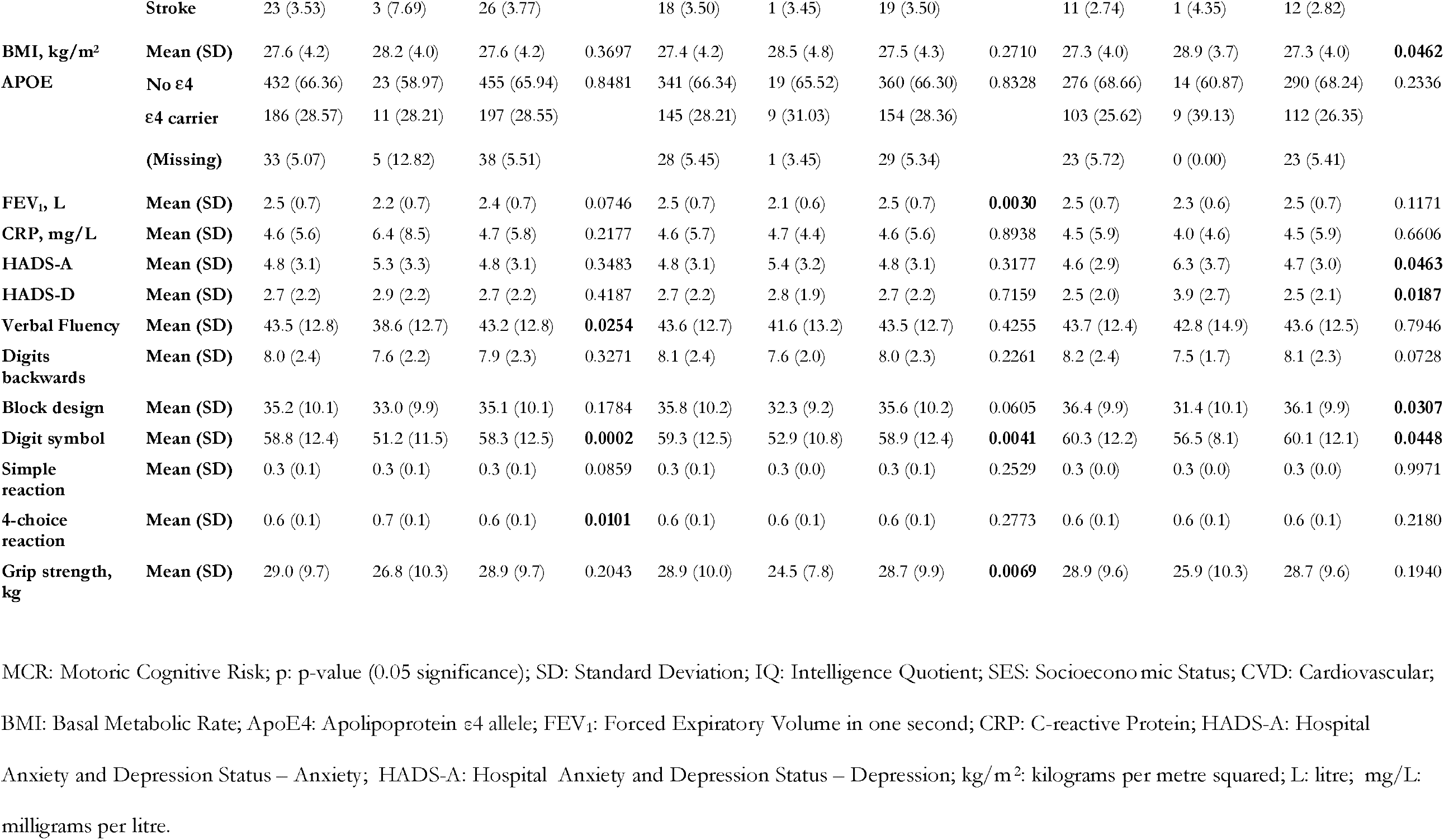
Baseline covariate differences of the participants according to MCR status at wave 3, wave 4 and wave 5

| Baseline Covariates |  | MCR at wave 3 (6 years follow-up) |  |  |  | MCR at wave 4 (9 years follow-up) |  |  |  | MCR at wave 5 (12 years follow-up) |  |  |  |
| --- | --- | --- | --- | --- | --- | --- | --- | --- | --- | --- | --- | --- | --- |
|  |  | No MCR | MCR | Total | p | No MCR | MCR | Total | p | No MCR | MCR | Total | p |
| Total N (%) |  | 651 (94.35) | 39 (5.65) | 690 |  | 514 (94.66) | 29 (5.34) | 543 |  | 402 (94.59) | 23 (5.41) | 425 |  |
| Age, years | Mean (SD) | 69.5 (0.8) | 69.9 (0.7) | 69.5 (0.8) | 0.0008 | 69.5 (0.8) | 69.8 (0.7) | 69.5 (0.8) | 0.0391 | 69.5 (0.8) | 69.5 (0.9) | 69.5 (0.8) | 0.9455 |
| Sex | Male | 340 (52.23) | 19 (48.72) | 359 (52.03) | 0.7423 | 260 (50.58) | 13 (44.83) | 273 (50.28) | 0.5725 | 200 (49.75) | 8 (34.78) | 208 (48.94) | 0.1999 |
|  | Female | 311 (47.77) | 20 (51.28) | 331 (47.97) |  | 254 (49.42) | 16 (55.17) | 270 (49.72) |  | 202 (50.25) | 15 (65.22) | 217 (51.06) |  |
| Education, years | Mean (SD) | 10.8 (1.1) | 10.6 (1.1) | 10.8 (1.1) | 0.3414 | 10.9 (1.2) | 10.6 (1.2) | 10.9 (1.2) | 0.2088 | 10.9 (1.2) | 10.5 (1.2) | 10.9 (1.2) | 0.0837 |
| Age 11 IQ | Mean (SD) | 101.6 (15.4) | 100.8 (12.2) | 101.6 (15.3) | 0.7119 | 101.9 (15.3) | 101.7 (15.2) | 101.9 (15.2) | 0.9497 | 102.5 (15.0) | 103.5 (13.2) | 102.5 (14.9) | 0.7198 |
| SES | Non-manual | 520 (79.88) | 23 (58.97) | 543 (78.70) | 0.0056 | 417 (81.13) | 22 (75.86) | 439 (80.85) | 0.3269 | 335 (83.33) | 19 (82.61) | 354 (83.29) | 0.7659 |
|  | Manual | 123 (18.89) | 15 (38.46) | 138 (20.00) |  | 88 (17.12) | 7 (24.14) | 95 (17.50) |  | 60 (14.93) | 4 (17.39) | 64 (15.06) |  |
|  | (Missing) | 8 (1.23) | 1 (2.56) | 9 (1.30) |  | 9 (1.75) | 0 (0.00) | 9 (1.66) |  | 7 (1.74) | 0 (0.00) | 7 (1.65) |  |
| Marital status | Married | 465 (71.43) | 32 (82.05) | 497 (72.03) | 0.1981 | 371 (72.18) | 17 (58.62) | 388 (71.45) | 0.1384 | 288 (71.64) | 16 (69.57) | 304 (71.53) | 0.8149 |
|  | Not married | 186 (28.57) | 7 (17.95) | 193 (27.97) |  | 143 (27.82) | 12 (41.38) | 155 (28.55) |  | 114 (28.36) | 7 (30.43) | 121 (28.47) |  |
| Smoking status | Never | 325 (49.92) | 22 (56.41) | 347 (50.29) | 0.1391 | 277 (53.89) | 17 (58.62) | 294 (54.14) | 0.4597 | 209 (51.99) | 16 (69.57) | 225 (52.94) | 0.1784 |
|  | Ex-smoker | 283 (43.47) | 12 (30.77) | 295 (42.75) |  | 216 (42.02) | 10 (34.48) | 226 (41.62) |  | 179 (44.53) | 6 (26.09) | 185 (43.53) |  |
|  | Current | 43 (6.61) | 5 (12.82) | 48 (6.96) |  | 21 (4.09) | 2 (6.90) | 23 (4.24) |  | 14 (3.48) | 1 (4.35) | 15 (3.53) |  |
| Alcohol, units/week | Mean (SD) | 10.5 (13.5) | 8.8 (9.2) | 10.4 (13.3) | 0.2708 | 6.0 (1.0 to 15.0) | 5.0 (0.5 to 13.0) | 6.0 (1.0 to 15.0) | 0.4764 | 7.0 (1.0 to 15.0) | 3.0 (0.8 to 10.5) | 6.0 (1.0 to 15.0) | 0.1894 |
| CVD history | No CVD | 502 (77.11) | 27 (69.23) | 529 (76.67) | 0.2481 | 394 (76.65) | 24 (82.76) | 418 (76.98) | 0.6498 | 318 (79.10) | 18 (78.26) | 336 (79.06) | 1.0000 |
|  | CVD | 149 (22.89) | 12 (30.77) | 161 (23.33) |  | 120 (23.35) | 5 (17.24) | 125 (23.02) |  | 84 (20.90) | 5 (21.74) | 89 (20.94) |  |
| Stroke history | No stroke | 628 (96.47) | 36 (92.31) | 664 (96.23) | 0.1767 | 496 (96.50) | 28 (96.55) | 524 (96.50) | 1.0000 | 391 (97.26) | 22 (95.65) | 413 (97.18) | 0.4917 |
|  | **Stroke** | 23 (3.53) | 3 (7.69) | 26 (3.77) |  | 18 (3.50) | 1 (3.45) | 19 (3.50) |  | 11 (2.74) | 1 (4.35) | 12 (2.82) |  |
| **BMI, kg/m<sup>2</sup>** | **Mean (SD)** | 27.6 (4.2) | 28.2 (4.0) | 27.6 (4.2) | 0.3697 | 27.4 (4.2) | 28.5 (4.8) | 27.5 (4.3) | 0.2710 | 27.3 (4.0) | 28.9 (3.7) | 27.3 (4.0) | **0.0462** |
| **APOE** | **No ε4** | 432 (66.36) | 23 (58.97) | 455 (65.94) | 0.8481 | 341 (66.34) | 19 (65.52) | 360 (66.30) | 0.8328 | 276 (68.66) | 14 (60.87) | 290 (68.24) | 0.2336 |
|  | **ε4 carrier** | 186 (28.57) | 11 (28.21) | 197 (28.55) |  | 145 (28.21) | 9 (31.03) | 154 (28.36) |  | 103 (25.62) | 9 (39.13) | 112 (26.35) |  |
|  | **(Missing)** | 33 (5.07) | 5 (12.82) | 38 (5.51) |  | 28 (5.45) | 1 (3.45) | 29 (5.34) |  | 23 (5.72) | 0 (0.00) | 23 (5.41) |  |
| **FEV<sub>1</sub>, L** | **Mean (SD)** | 2.5 (0.7) | 2.2 (0.7) | 2.4 (0.7) | 0.0746 | 2.5 (0.7) | 2.1 (0.6) | 2.5 (0.7) | **0.0030** | 2.5 (0.7) | 2.3 (0.6) | 2.5 (0.7) | 0.1171 |
| **CRP, mg/L** | **Mean (SD)** | 4.6 (5.6) | 6.4 (8.5) | 4.7 (5.8) | 0.2177 | 4.6 (5.7) | 4.7 (4.4) | 4.6 (5.6) | 0.8938 | 4.5 (5.9) | 4.0 (4.6) | 4.5 (5.9) | 0.6606 |
| **HADS-A** | **Mean (SD)** | 4.8 (3.1) | 5.3 (3.3) | 4.8 (3.1) | 0.3483 | 4.8 (3.1) | 5.4 (3.2) | 4.8 (3.1) | 0.3177 | 4.6 (2.9) | 6.3 (3.7) | 4.7 (3.0) | **0.0463** |
| **HADS-D** | **Mean (SD)** | 2.7 (2.2) | 2.9 (2.2) | 2.7 (2.2) | 0.4187 | 2.7 (2.2) | 2.8 (1.9) | 2.7 (2.2) | 0.7159 | 2.5 (2.0) | 3.9 (2.7) | 2.5 (2.1) | **0.0187** |
| **Verbal Fluency** | **Mean (SD)** | 43.5 (12.8) | 38.6 (12.7) | 43.2 (12.8) | **0.0254** | 43.6 (12.7) | 41.6 (13.2) | 43.5 (12.7) | 0.4255 | 43.7 (12.4) | 42.8 (14.9) | 43.6 (12.5) | 0.7946 |
| **Digits backwards** | **Mean (SD)** | 8.0 (2.4) | 7.6 (2.2) | 7.9 (2.3) | 0.3271 | 8.1 (2.4) | 7.6 (2.0) | 8.0 (2.3) | 0.2261 | 8.2 (2.4) | 7.5 (1.7) | 8.1 (2.3) | 0.0728 |
| **Block design** | **Mean (SD)** | 35.2 (10.1) | 33.0 (9.9) | 35.1 (10.1) | 0.1784 | 35.8 (10.2) | 32.3 (9.2) | 35.6 (10.2) | 0.0605 | 36.4 (9.9) | 31.4 (10.1) | 36.1 (9.9) | **0.0307** |
| **Digit symbol** | **Mean (SD)** | 58.8 (12.4) | 51.2 (11.5) | 58.3 (12.5) | **0.0002** | 59.3 (12.5) | 52.9 (10.8) | 58.9 (12.4) | **0.0041** | 60.3 (12.2) | 56.5 (8.1) | 60.1 (12.1) | **0.0448** |
| **Simple reaction** | **Mean (SD)** | 0.3 (0.1) | 0.3 (0.1) | 0.3 (0.1) | 0.0859 | 0.3 (0.1) | 0.3 (0.0) | 0.3 (0.1) | 0.2529 | 0.3 (0.0) | 0.3 (0.0) | 0.3 (0.0) | 0.9971 |
| **4-choice reaction** | **Mean (SD)** | 0.6 (0.1) | 0.7 (0.1) | 0.6 (0.1) | **0.0101** | 0.6 (0.1) | 0.6 (0.1) | 0.6 (0.1) | 0.2773 | 0.6 (0.1) | 0.6 (0.1) | 0.6 (0.1) | 0.2180 |
| **Grip strength, kg** | **Mean (SD)** | 29.0 (9.7) | 26.8 (10.3) | 28.9 (9.7) | 0.2043 | 28.9 (10.0) | 24.5 (7.8) | 28.7 (9.9) | **0.0069** | 28.9 (9.6) | 25.9 (10.3) | 28.7 (9.6) | 0.1940 |

MCR: Motoric Cognitive Risk; p: p-value (0.05 significance); SD: Standard Deviation; IQ: Intelligence Quotient; SES: Socioeconomic Status; CVD: Cardiovascular;
BMI: Basal Metabolic Rate; ApoE4: Apolipoprotein ε4 allele; FEV<sub>1</sub>: Forced Expiratory Volume in one second; CRP: C-reactive Protein; HADS-A: Hospital
Anxiety and Depression Status – Anxiety; HADS-A: Hospital Anxiety and Depression Status – Depression; kg/m<sup>2</sup>: kilograms per metre squared; L: litre; mg/L:
milligrams per litre.

### Subgroup analysis

We performed a sensitivity analysis of the same baseline covariate differences according to MCR status of withdrawers before wave 4 (Supplementary Table 2) and wave 5 (Supplementary Table 3) to assess for selection bias due to attrition. We used an identical statistical approach as for the main analysis. Only verbal fluency at wave 3 (p = 0.015) and FEV_1_ (p = 0.0079) at wave 4 differed significantly between the non-MCR and MCR groups.

### MCR, MCI and Frailty level overlap

The overlap between MCR, MCI, Prefrailty, and Frailty is presented in Figure 2. MCI was derived at waves 3, 4 and 5 but Frailty level was only derived at waves 4 and 5 due to the unavailability of necessary variables at wave 3. As a proportion of those participants with either MCR or MCI, the overlap between MCR and MCI is remarkably consistent across each wave – 10.6% at wave 3, 11.6% at wave 4, and 10.4% at wave 5, averaging 10.9% (95% CI 7.4-15.2) across all three waves. Of those participants with MCR, overlap with MCI is 39.3% at wave 3, 52.6% at wave 4, and 50% at wave 5, averaging 46% (95% CI 33.4-59.1) across all three waves. Once Frailty and Prefrailty are added, only one participant with MCR does not have either Prefrailty or Frailty at waves 4 and 5.

**Figure 2:**
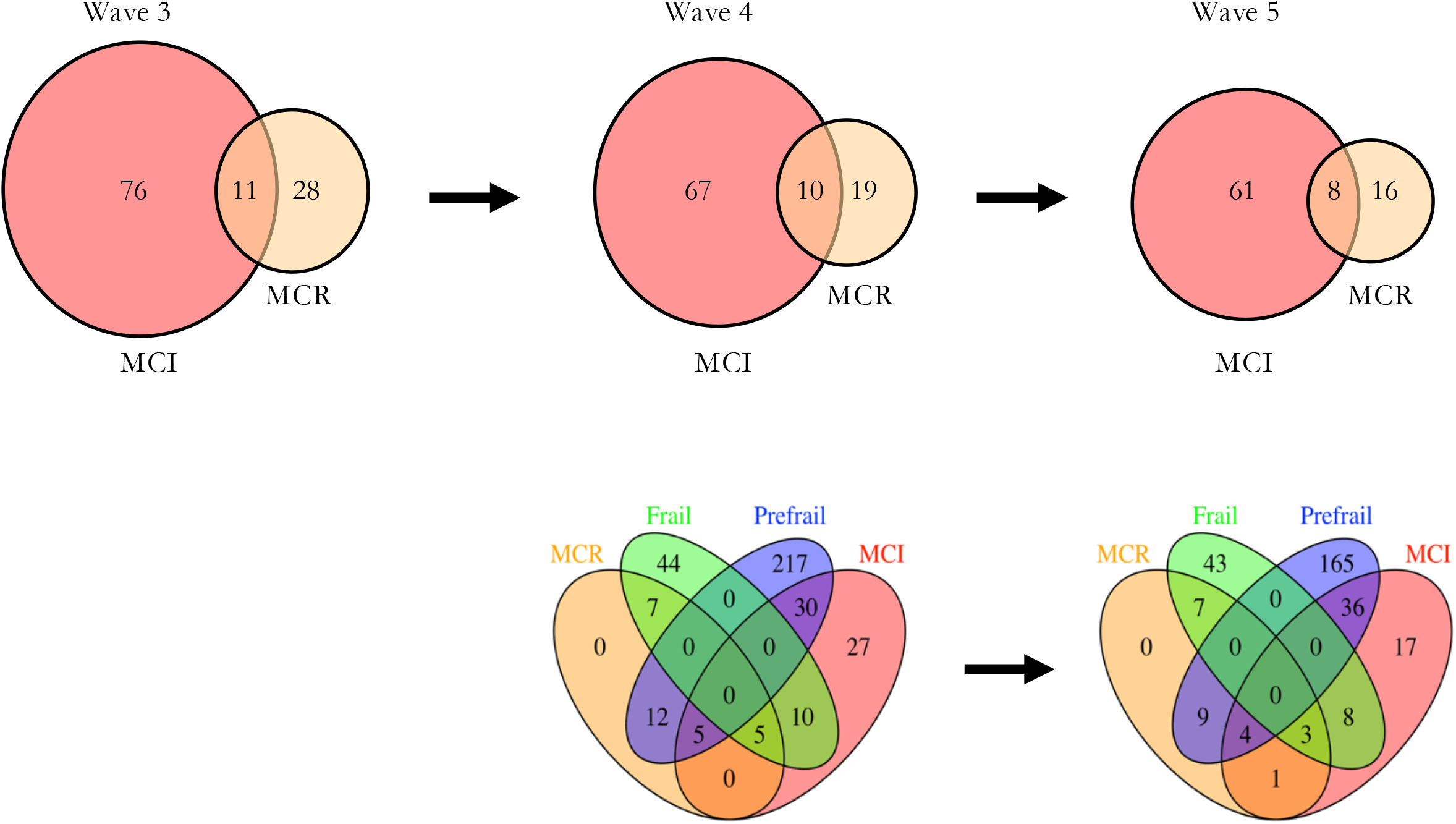
The overlap between MCR, MCI and Frailty level in the LBC1936 cohort. *Top*, MCR and MCI overlap at waves 3, 4 and 5. *Bottom*, MCR, MCI, Frailty and Prefrailty overlap at waves 4 and 5. Frailty and Prefrailty were not measured at wave 3 due to lack of necessary variables. By definition, participants cannot be classed as Frail and Prefrail simultaneously.

## Discussion

### MCR prevalence

In this cohort of older Scottish adults, we have determined the prevalence of MCR syndrome, described associated risk factors and assessed its overlap with Mild Cognitive Impairment, Prefrailty, and Frailty. The prevalence of MCR averaged 5.5% (95% CI 4.5-6.7) over three waves over a six-year period. There was no significant difference between men and women. This prevalence is lower than the 9.7% pooled rate of 22 cohorts from 17 countries, mean age 73.6 years (±8.2).^19^ This may be partly explained by attrition bias, as the MCR prevalence of withdrawers at waves 3 and 4 is higher than completers. Interestingly though, two studies of our closest neighbours, The English Longitudinal Study of Ageing^19^ and The Irish Longitudinal Study on Ageing^20^ reported rates of 2% and 2.6% respectively. LBC1936 MCR prevalence may be higher than those of the English and Irish cohorts due to the older average age of the LBC1936 participants and the accompanying increased rates of cognitive complaints.

The gait speed cut-offs in our study were higher in men than women, and lower with older age, in keeping with the literature.^21^ Our gait speed cut-offs were higher than most reported in other studies of MCR for each age- and sex-matched group.^2,4,22^ In fact, the slow gait cut-offs in LBC1936 were similar to the mean usual gait speeds for similar groupings in a comprehensive meta-analysis of usual gait speeds of 23,111 individuals from 12 countries.^21^ This could indicate that the average usual walking speed is quite fast in Scotland but more likely reflects the level of health in LBC1936 participants. Without published national reference age- and sex-matched gait speeds, it is difficult to be sure.

### Baseline covariate differences

Interestingly, despite the narrow age range of LBC1936 participants, we noted a significant association between older age and the presence of MCR in two of the three waves. This mixed picture is in keeping with a recent meta-analysis of factors associated with MCR, which found that the majority but not all of the 22 studies reported age as an associated factor for the presence of MCR.^12^

Lower socioeconomic status, as defined by having had a manual occupation, was associated with having MCR later in life. At first, this seems counterintuitive as one could hypothesize that keeping physically active at work would reduce gait speed slowing in later life.^23^ However, it is probable that other factors associated at a population level with having a manual job, such as less years in formal education, diminish any protection offered from being physically active at work.

It is perhaps unsurprising that MCR was consistently associated with poorer scores in tests of executive function across the three waves as slow gait speed has been repeatedly associated with these tests in the literature.^24^ One hypothesis is that walking requires significant top-down coordination and planning as well as attention and response inhibition, particularly when walking in an unfamiliar environment.^24^ Indeed, imaging studies have shown that the brain areas most responsible for executive function tasks are often more damaged in the MCR group than the non-MCR group.^5,25,26^ In particular, the digit symbol test was the only covariate to remain significant across all three waves (wave 3 *p* = 0.0002, wave 4 *p* = 0.0041, wave 5 *p* = 0.0448), highlighting it as an especially sensitive marker of MCR. The digit symbol test, a subtest from the Wechsler Adult Intelligence Scale-III UK,^27^ predominantly assesses processing speed. The participant enters a symbol according to a given number-symbol code, completing as many as possible in two minutes. This test has been previously found to serve as a biomarker of risk of clinical disorders of cognition and mobility.^28^

This is the first study to examine the association between early-life intelligence test score and MCR status later in life. There was no significant relationship found. This is an important finding as it does not support previous work detailing an association between lower early life intelligence scores and slower gait and poorer cognitive performance.^14^ Consistent with the literature, alcohol consumption was also not significantly associated with MCR status.^6,12,13^ More surprisingly, however, were the findings that years of education, BMI, stroke, and cardiovascular disease were not associated with MCR status. These covariates have generally been associated with MCR.^6,12,13^ This variation may be because the LBC1936 cohort consists of participants from an affluent area of Scotland who volunteered to take part, and the average years of education, as well as general physical fitness, is notably higher than the general population.^8,9^ This healthy volunteer bias is common to longitudinal studies of ageing and should be considered when deciding how generalisable our findings are to the clinical population. It may also be that our study did not have significant power to detect a significant relationship, an idea supported by our effect sizes which, although not significant, are generally in the same direction as larger studies.^6,12,13^

### Subgroup analysis

Our sensitivity analysis comparing the baseline covariates differences according to MCR status of withdrawers before wave 4 and wave 5 was reassuring as there were very few significant differences. However, the rate of withdrawal for individuals with MCR was significantly higher than individuals without MCR, indicating likely selection bias due to attrition. Attrition from ill-health or mortality is a common and often unavoidable bias of longitudinal studies of ageing.

### Overlap of MCR, MCI, Prefrailty and Frailty

The level of overlap between MCR and MCI shows that these two concepts, although derived using similar criteria and thus sharing some participants, also capture different cohorts of people. Our findings reinforce previous research which determined that we should view MCR as complementary to MCI rather than replacing it.^2^ Assessing for both prodromes is likely to yield more people at high risk of developing dementia.^2^ This is the first study to explore the overlap of MCR and Frailty according to Fried phenotype. Considering the make-up of the Fried phenotype, it is entirely unsurprising that almost all the MCR group had either Prefrailty or Frailty. The decision as to which prodrome state is more clinically useful will ultimately be determined by balancing the cost, effort and time taken to measure each prodrome with the prognostic value for the outcome in question.

## Conclusion

Prevalence rates of MCR in this Scottish cohort are lower than the global average but higher than in neighbouring countries. Future Lothian Birth Cohort 1936 research should assess the risk factors associated with MCR to validate previous findings and analyse novel predictive factors. Work exploring the association between socioeconomic status and MCR could help address the disparities in health care and health outcomes in the United Kingdom. Examining the prognostic value of MCR as a predictor of cognitive decline, specifically executive function, in LBC1936 is a vital avenue to explore. Our study can serve as a foundation for future studies to improve dementia risk assessments and potentially develop new interventions to reduce incident dementia.

## Data Availability

All data produced in the present study are available upon reasonable request to the authors

