## Supplementary file for "Prevalence and predictors of Motoric Cognitive Risk syndrome in a community-dwelling older Scottish population: a longitudinal observational study"

Supplementary Table 1: Measurement and derivation of covariates of note

**Body Mass Index**

Weight in kg/Height in m^2^

**Age 11 intelligence quotient (IQ)**

Based on LBC1936 participants’ scores in the Moray House Test at age 11.^8^ Raw scores were corrected for age in days at the time of testing and converted to an IQ scale where mean (SD) = 100 (15).

**Socioeconomic status**

Based on occupational social class (professional, managerial, nonmanual skilled, manual skilled, manual semiskilled, unskilled), as coded at the 1980 census.^29^ Married females were assigned a social class based on their husband’s occupation if that was higher than their own. We collapsed these six categories into either nonmanual or manual, with professional, managerial, and nonmanual skilled being classed as nonmanual and manual skilled, manual semiskilled, and unskilled being classed as manual. This improved the distribution of participants between categories and the manual/nonmanual association is important to explore when examining the MCR phenotype which includes a significant manual or ‘motoric’ component.

**Mild Cognitive Impairment**

As defined by the National Institute on Aging-Alzheimer’s Association (NIA-AA) workgroups on diagnostic guidelines for Alzheimer disease. MCI was recently derived in LBC1936 using very similar criteria to MCR,^15^ except that MCI uses an objective impairment in one or more cognitive domains criterion instead of an objective slow gait criterion.

**Physical Frailty**

As determined by the Fried phenotype. This was recently derived in LBC1936 using five pre-specified criteria: weight loss, exhaustion, physical health, walking speed, and grip strength.^16^ The presence of one or two of these dimensions indicated that an individual is Pre-frail, whilst three or more indicated Frailty.

**Grip Strength**

This was assessed using a Jamar Hydraulic Hand Dynamometer. It was administered on a one-to-one basis. At each assessment, it was tested three times in both hands, and the best recording (in kilograms) from each hand was recorded. We combined both left- and right-hand measures to create a single combined average of both hands.

Supplementary Table 2: Withdrawers with MCR at wave 3 & explanatory variables at wave 1

| Dependent: MCR_w3 |  | No MCR | MCR | Total | p |
| --- | --- | --- | --- | --- | --- |
| Total N (%) |  | 246 (91.45) | 23 (8.55) | 269 |  |
| **Age, years** | Mean (SD) | 69.5 (0.8) | 69.8 (0.7) | 69.6 (0.8) | 0.1841 |
| **Sex** | Male | 141 (57.32) | 11 (47.83) | 152 (56.51) | 0.3891 |
|  | Female | 105 (42.68) | 12 (52.17) | 117 (43.49) |  |
| **Education, years** | Mean (SD) | 10.7 (1.1) | 10.6 (1.1) | 10.6 (1.1) | 0.8652 |
| **Age 11 IQ** | Mean (SD) | 99.8 (16.1) | 102.6 (11.4) | 100.0 (15.8) | 0.3229 |
| **SES** | Non-manual | 179 (72.76) | 13 (56.52) | 192 (71.38) | 0.0939 |
|  | Manual | 65 (26.42) | 10 (43.48) | 75 (27.88) |  |
|  | (Missing) | 2 (0.81) | 0 (0.00) | 2 (0.74) |  |
| **Marital status** | Married | 180 (73.17) | 19 (82.61) | 199 (73.98) | 0.4571 |
|  | Not married | 66 (26.83) | 4 (17.39) | 70 (26.02) |  |
| **Smoking status** | Never | 113 (45.93) | 12 (52.17) | 125 (46.47) | 0.8655 |
|  | Ex-smoker | 102 (41.46) | 9 (39.13) | 111 (41.26) |  |
|  | Current | 31 (12.60) | 2 (8.70) | 33 (12.27) |  |
| **Alcohol, units/week** | Median (IQR) | 6.0 (0.5 to 15.0) | 7.0 (2.0 to 16.8) | 6.0 (0.5 to 15.0) | 0.7522 |
| **CVD history** | No CVD | 180 (73.17) | 17 (73.91) | 197 (73.23) | 1.0000 |
|  | CVD | 66 (26.83) | 6 (26.09) | 72 (26.77) |  |
| **Stroke history** | No stroke | 231 (93.90) | 23 (100.00) | 254 (94.42) | 0.6254 |
|  | Stroke | 15 (6.10) | 0 (0.00) | 15 (5.58) |  |
| **BMI** | Mean (SD) | 27.9 (4.6) | 28.6 (4.8) | 28.0 (4.6) | 0.4976 |
| **APOE** | No ε4 | 158 (64.23) | 11 (47.83) | 169 (62.83) | 0.4517 |
|  | ε4 carrier | 77 (31.30) | 8 (34.78) | 85 (31.60) |  |
|  | (Missing) | 11 (4.47) | 4 (17.39) | 15 (5.58) |  |
| **FEV_1_, L** | Mean (SD) | 2.4 (0.7) | 2.2 (0.6) | 2.4 (0.7) | 0.1162 |
| **CRP, mg/L** | Mean (SD) | 4.9 (4.9) | 7.4 (10.4) | 5.1 (5.6) | 0.2868 |
| **HADS-A** |  |  |  |  |  |
| **HADS-D** | Mean (SD) | 2.9 (2.3) | 2.8 (2.0) | 2.9 (2.3) | 0.7957 |
| **Verbal Fluency** | Mean (SD) | 43.0 (13.5) | 37.5 (9.5) | 42.6 (13.3) | **0.0157** |
| **Digits backwards** | Mean (SD) | 7.6 (2.3) | 7.8 (2.3) | 7.6 (2.3) | 0.6247 |
| **Block design** | Mean (SD) | 33.5 (10.1) | 34.3 (11.1) | 33.6 (10.2) | 0.7363 |
| **Digit symbol** | Mean (SD) | 56.0 (12.7) | 51.8 (13.0) | 55.6 (12.7) | 0.1549 |
| **Simple reaction** | Mean (SD) | 0.3 (0.1) | 0.3 (0.0) | 0.3 (0.1) | 0.9319 |
| **4-choice reaction** |  |  |  |  |  |
| **Grip strength, kg** | Mean (SD) | 30.1 (10.3) | 27.1 (8.5) | 29.9 (10.2) | 0.1168 |

Supplementary Table 3: Withdrawers with MCR at wave 4 & explanatory variables at wave 1

| Dependent: MCR_w4 |  | No MCR | MCR | Total | p |
| --- | --- | --- | --- | --- | --- |
| Total N (%) |  | 113 (88.98) | 14 (11.02) | 127 |  |
| **Age, years** | Mean (SD) | 69.6 (0.8) | 70.0 (0.6) | 69.7 (0.8) | 0.0577 |
| **Sex** | Male | 63 (55.75) | 6 (42.86) | 69 (54.33) | 0.4044 |
|  | Female | 50 (44.25) | 8 (57.14) | 58 (45.67) |  |
| **Education, years** | Mean (SD) | 10.8 (1.2) | 10.6 (1.3) | 10.7 (1.2) | 0.6365 |
| **Age 11 IQ** | Mean (SD) | 99.9 (16.0) | 99.8 (18.9) | 99.9 (16.2) | 0.9737 |
| **SES** | Non-manual | 81 (71.68) | 10 (71.43) | 91 (71.65) | 1.0000 |
|  | Manual | 30 (26.55) | 4 (28.57) | 34 (26.77) |  |
|  | (Missing) | 2 (1.77) | 0 (0.00) | 2 (1.57) |  |
| **Marital status** | Married | 82 (72.57) | 9 (64.29) | 91 (71.65) | 0.5374 |
|  | Not married | 31 (27.43) | 5 (35.71) | 36 (28.35) |  |
| **Smoking status** | Never | 63 (55.75) | 8 (57.14) | 71 (55.91) | 1.0000 |
|  | Ex-smoker | 42 (37.17) | 5 (35.71) | 47 (37.01) |  |
|  | Current | 8 (7.08) | 1 (7.14) | 9 (7.09) |  |
| **Alcohol, units/week** | Median (IQR) | 5.0 (0.5 to 17.0) | 7.5 (1.6 to 13.8) | 5.0 (0.5 to 15.5) | 0.9691 |
| **CVD history** | No CVD | 81 (71.68) | 11 (78.57) | 92 (72.44) | 0.7562 |
|  | CVD | 32 (28.32) | 3 (21.43) | 35 (27.56) |  |
| **Stroke history** | No stroke | 106 (93.81) | 13 (92.86) | 119 (93.70) | 1.0000 |
|  | Stroke | 7 (6.19) | 1 (7.14) | 8 (6.30) |  |
| **BMI** | Mean (SD) | 28.0 (5.2) | 27.0 (3.6) | 27.9 (5.1) | 0.3450 |
| **APOE** | No e4 | 66 (58.41) | 9 (64.29) | 75 (59.06) | 0.7643 |
|  | e4 carrier | 42 (37.17) | 4 (28.57) | 46 (36.22) |  |
|  | (Missing) | 5 (4.42) | 1 (7.14) | 6 (4.72) |  |
| **FEV_1_, L** | Mean (SD) | 2.4 (0.7) | 2.0 (0.5) | 2.4 (0.7) | **0.0079** |
| **CRP, mg/L** | Mean (SD) | 4.7 (4.2) | 5.7 (5.4) | 4.8 (4.4) | 0.5537 |
| **HADS-A** |  |  |  |  |  |
| **HADS-D** | Mean (SD) | 3.2 (2.7) | 2.7 (1.4) | 3.2 (2.6) | 0.2822 |
| **Verbal Fluency** | Mean (SD) | 43.4 (13.8) | 40.9 (12.7) | 43.1 (13.6) | 0.5123 |
| **Digits backwards** | Mean (SD) | 7.7 (2.4) | 7.3 (1.6) | 7.6 (2.3) | 0.4520 |
| **Block design** | Mean (SD) | 34.5 (10.6) | 30.4 (9.0) | 34.0 (10.5) | 0.1392 |
| **Digit symbol** | Mean (SD) | 55.9 (13.3) | 51.0 (12.5) | 55.3 (13.3) | 0.1892 |
| **Simple reaction** | Mean (SD) | 0.3 (0.1) | 0.3 (0.0) | 0.3 (0.1) | 0.3940 |
| **4-choice reaction** |  |  |  |  |  |
| **Grip strength, kg** | Mean (SD) | 30.0 (11.0) | 24.4 (9.3) | 29.4 (10.9) | 0.0539 |
